# Assessing Retrieval-Augmented Large Language Model Performance in Emergency Department ICD-10-CM Coding Compared to Human Coders

**DOI:** 10.1101/2024.10.15.24315526

**Authors:** Eyal Klang, Idit Tessler, Donald U Apakama, Ethan Abbott, Benjamin S Glicksberg, Monique Arnold, Akini Moses, Ankit Sakhuja, Ali Soroush, Alexander W Charney, David L. Reich, Jolion McGreevy, Nicholas Gavin, Brendan Carr, Robert Freeman, Girish N Nadkarni

## Abstract

**Background:** Accurate medical coding is essential for clinical and administrative purposes but complicated, time-consuming, and biased. This study compares Retrieval-Augmented Generation (RAG)-enhanced LLMs to provider-assigned codes in producing ICD-10-CM codes from emergency department (ED) clinical records.

**Methods:** Retrospective cohort study using 500 ED visits randomly selected from the Mount Sinai Health System between January and April 2024. The RAG system integrated past 1,038,066 ED visits data (2021-2023) into the LLMs’ predictions to improve coding accuracy. Nine commercial and open-source LLMs were evaluated. The primary outcome was a head-to-head comparison of the ICD-10-CM codes generated by the RAG-enhanced LLMs and those assigned by the original providers. A panel of four physicians and two LLMs blindly reviewed the codes, comparing the RAG-enhanced LLM and provider-assigned codes on accuracy and specificity.

**Findings:** RAG-enhanced LLMs demonstrated superior performance to provider coders in both the accuracy and specificity of code assignments. In a targeted evaluation of 200 cases where discrepancies existed between GPT-4 and provider-assigned codes, human reviewers favored GPT-4 for accuracy in 447 instances, compared to 277 instances where providers’ codes were preferred (p<0.001). Similarly, GPT-4 was selected for its superior specificity in 509 cases, whereas human coders were preferred in only 181 cases (p<0.001). Smaller open-access models, such as Llama-3.1-70B, also demonstrated substantial scalability when enhanced with RAG, with 218 instances of accuracy preference compared to 90 for providers’ codes. Furthermore, across all models, the exact match rate between LLM-generated and provider-assigned codes significantly improved following RAG integration, with Qwen-2-7B increasing from 0.8% to 17.6% and Gemma-2-9b-it improving from 7.2% to 26.4%.

**Interpretation:** RAG-enhanced LLMs improve medical coding accuracy in EDs, suggesting clinical workflow applications. These findings show that generative AI can improve clinical outcomes and reduce administrative burdens.

**Funding:** This work was supported in part through the computational and data resources and staff expertise provided by Scientific Computing and Data at the Icahn School of Medicine at Mount Sinai and supported by the Clinical and Translational Science Awards (CTSA) grant UL1TR004419 from the National Center for Advancing Translational Sciences. Research reported in this publication was also supported by the Office of Research Infrastructure of the National Institutes of Health under award number S10OD026880 and S10OD030463. The content is solely the responsibility of the authors and does not necessarily represent the official views of the National Institutes of Health. The funders played no role in study design, data collection, analysis and interpretation of data, or the writing of this manuscript.

**Twitter Summary:** A study showed AI models with retrieval-augmented generation outperformed human doctors in ED diagnostic coding accuracy and specificity. Even smaller AI models perform favorably when using RAG. This suggests potential for reducing administrative burden in healthcare, improving coding efficiency, and enhancing clinical documentation.

## Introduction

Utilization by the International Classification of Diseases (ICD) system is a key requirement for clinical, financial and administration purposes, including clinical record keeping, public health monitoring, scholarly research, and medical reimbursement. ^1–3^ A high level of accuracy is essential to meet these goals. Current ICD coding systems, despite their importance in clinical and administrative tasks, are time-consuming and prone to errors, resulting in incorrect or incomplete documentation.

Efforts to automate the extraction of medical codes from clinical texts primarily use large language models (LLMs). Nonetheless, previous research, including our own,^3^ indicates that LLMs tend to produce inaccurate mappings when presented solely with code descriptions, often resulting in incorrect or hallucinated codes.^4,5^ Retrieval-Augmented Generation (RAG) integrates the generative capabilities of LLMs with a retrieval component that supplements the generation process with relevant external, verifiable data. ^6^ In this framework, a smaller LLM performs a semantic similarity search to retrieve relevant information, which then informs the generation process. This method not only supplements but significantly enhances the accuracy of the LLM’s outputs by using appropriate historical data points. ^7,8^ Previous applications of RAG in predicting emergency department admissions and patient triage have underscored its efficacy in improving model performance. ^9,10^

Given the mixed outcomes in previous LLM applications to medical coding and the potential seen in RAG-enhanced models to improve the efficacy and accuracy of clinical tasks, this study aims to evaluate the accuracy of RAG-enhanced LLMs in assigning primary diagnoses to patients discharged from emergency departments and to directly compare these AI-generated codes with those determined by ED providers.

## Methods

### Study Design

This retrospective cohort study analyzed the performance of a LLM with RAG system in generating ICD-10-CM codes and descriptions from clinical notes of patients discharged home from the ED. We specifically focused on patients discharged home because ICD codes for admitted patients may change as more information becomes available during their hospital stay. The study utilized a random sample of patients from the Mount Sinai Health System (MSHS), spanning multiple hospital emergency departments. Institutional Review Board (IRB) approval was obtained, and informed consent was waived by the IRB committee.

### Participants

We selected 500 adult patient emergency department (ED) visits from six Mount Sinai Health System (MSHS) hospitals. These visits, dated between January 1, 2024, and April 30, 2024, were chosen randomly from patients discharged home. Each record included the arrival and discharge dates, ED physician notes, and a final assigned ICD-10-CM primary diagnosis code. We retrieved original diagnosis ICD codes for all cases which were assigned in the MSHS EDs by attending clinicians during their standard documentation workflows.

### RAG Database

To assemble the RAG database for analysis, we collected data from all adult ED visits across the included MSHS hospitals from 2021 to 2023. This dataset included the primary diagnosis’s ICD code and code description. We calculated the frequency of each assigned ICD code during this period (i.e., the number of times a specific code was assigned divided by the total number of assigned codes).

### Development Phase

In the preliminary phase, we used 50 different ED discharge summaries w to iteratively refine the prompt structure for LLMs, optimizing their ICD code generation through systematic prompt testing.

### Data Collection

We retrieved data using Structured Query Language (SQL) queries to the MSHS Electronic Health Records (EHR) system (EPIC systems). All ED physician notes, written prior to patient discharge and code assignment, were concatenated into a single text for each patient. These texts served as inputs for the LLM predictions.

### Primary Outcome Measures

The primary outcome was a head-to-head comparison of the ICD-10-CM codes generated by the RAG-enhanced LLMs and those assigned by the original providers. A panel of four physicians and two LLMs reviewed the codes in a double-blind manner, comparing the RAG-enhanced LLM and provider-assigned codes to determine which was more accurate, more specific, or if both were equal.

### Experiment Procedure

Figure 1 outlines the experimental design, which includes the following stages:

1. <u>Initial Code Prediction Prompt:</u> Each of the nine evaluated LLMs predicted the primary ICD-10-CM diagnosis code and its description from clinical notes (See Supplementary Materials, Prompt 1).
2. <u>Retrieval-Augmented Generation (RAG):</u> This process involved querying a database of previous ED visits for ICD code descriptions to identify the ten most related codes. Each code description was also accompanied by its frequency of ED visits at MSHS between 2021 and 2023.
3. <u>Refinement Prompt:</u> The ten identified code descriptions were re-submitted to each of the nine LLMs along with the ED notes to select the most appropriate code description and its associated code (See Supplementary Materials, Prompt 2).
4. <u>LLM Evaluations:</u> Two large open-access LLMs (Llama-3.1-70B, Qwen-2-72B) were independently used to compare “head-to-head” all the LLM-assigned and human-assigned codes for accuracy and specificity. Each case was inputted with the case notes and both the provider-assigned and RAG-LLM-assigned code descriptions. Scores of 1 (human wins), 2 (LLM wins), or 3 (equal) were assigned for the accuracy and specificity categories (See Supplementary Materials, Prompt 3). The reviewing models were blinded to the origin of the code assignments to ensure unbiased evaluations. The reviewing models were also instructed to provide reasoning for their scoring decisions.
5. <u>Human Evaluation:</u> Four reviewers—two ED attendings and two ED residents—unaware of the origin of the code descriptions, evaluated the accuracy and specificity of a single LLM (GPT-4). They compared its code description assignments against the original provider-assigned code descriptions using the case notes for context. The reviewers assigned scores of 1 (human wins), 2 (LLM wins), or 3 (equal) for each category.

**Figure 1.**
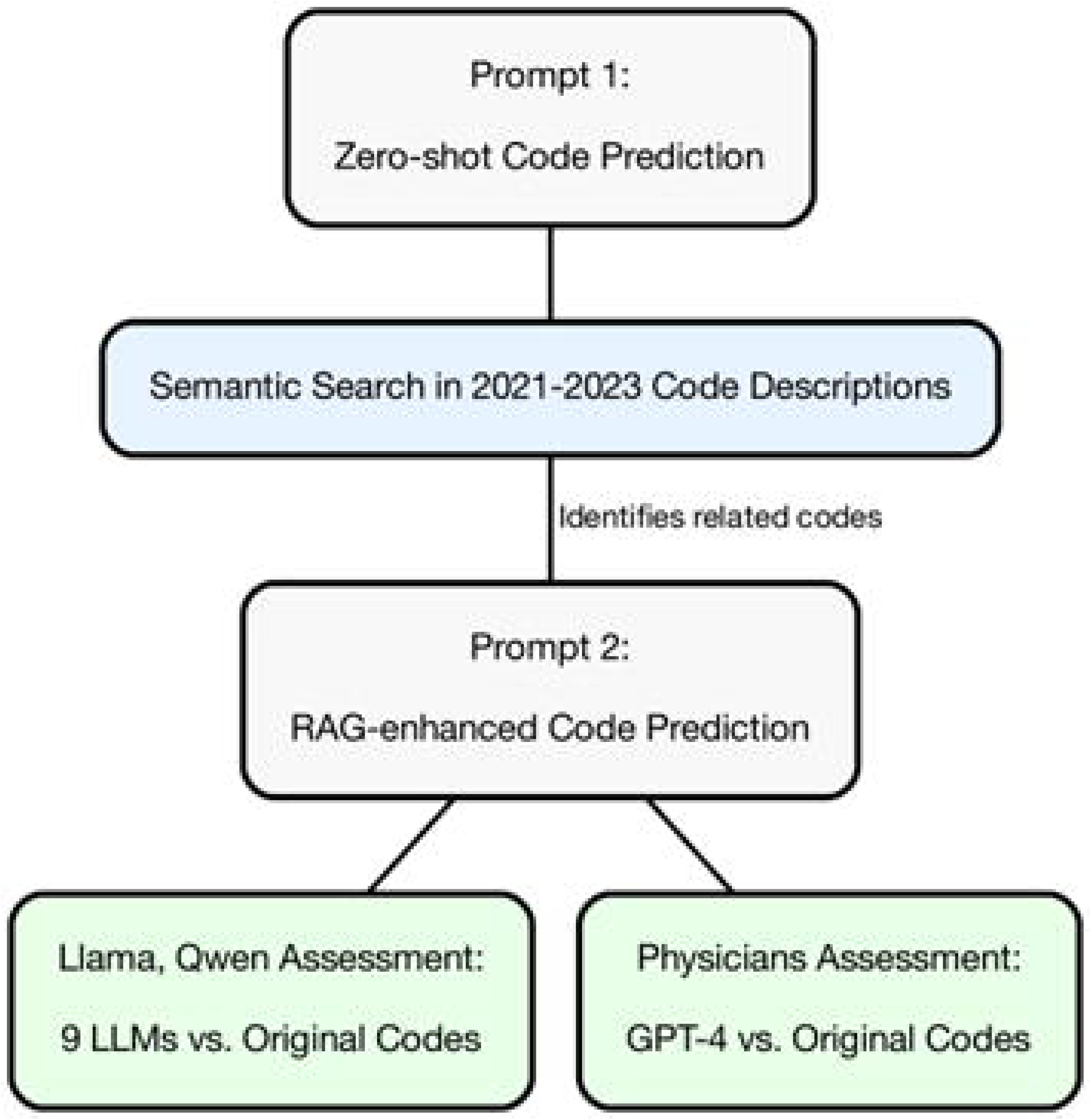
The study’s experimental design. Outlines of key steps in the research process, including initial code prediction, retrieval-augmented generation (RAG), refinement, and evaluation stages.

### ICD Coding Stages

⍰ <u>First Stage:</u> Each LLM was prompted up to three times to return a JSON object containing “code” and “description” items. If a valid JSON object was not returned after three attempts, it was marked as a “failed” JSON.
⍰ <u>Second Stage:</u> If the first stage was successful, each LLM was prompted again up to three times for a valid JSON. We also required that the returned ICD code be one of the ten RAG retrieved codes. If unsuccessful after three attempts, the ICD code from the first prompt was used instead for the second code assignment.

### Justification of Evaluation Criteria and Process

This study employs a direct comparison framework. The reviewers (LLMs and physicians) were asked to compare human vs. LLM code assignments based on their *accuracy* and *specificity*. These terms were selected for their clear, universally understood meanings, facilitating a straightforward yet effective evaluation process.

Accuracy measures whether the codes correctly reflect the conditions documented in the clinical notes. Specificity assesses the precision of these codes in detailing the specific conditions without generalization. The simplicity of these terms ensures that all judges, both human physicians and AI models, can apply them consistently and objectively, following the clear instructions provided.

Our methodological approach is designed to offer clear, unambiguous benchmarking of LLMs against human performance. This head-to-head comparison method scored as 1 for human superiority, 2 for LLM superiority, and 3 for equivalent performance, minimizes subjective interpretation and focuses solely on the factual correctness and relevance of the code assignments.

### Language Models Evaluated

The nine LLMs used in this study are presented in Table 1. All the open access models were of the “Instruct” version. Access to the GPT models was provided through the MSHS Azure tenant environment using the OpenAI API. Llama, Qwen, Gemma and Phi models were accessed via a 4xH100 80GB local MSH cluster through the Hugging Face interface. Default model settings were used for all the models.

**Table 1.**
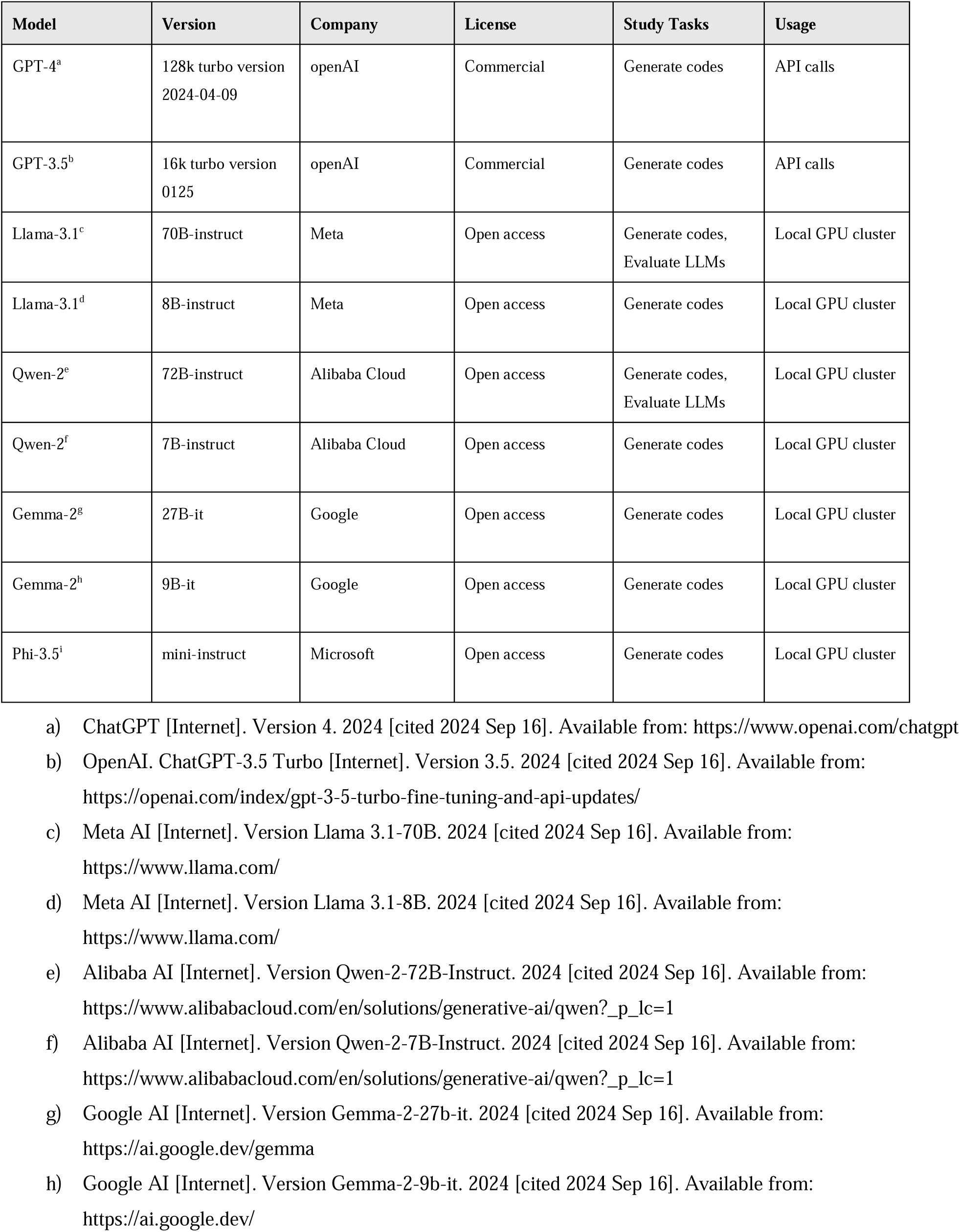
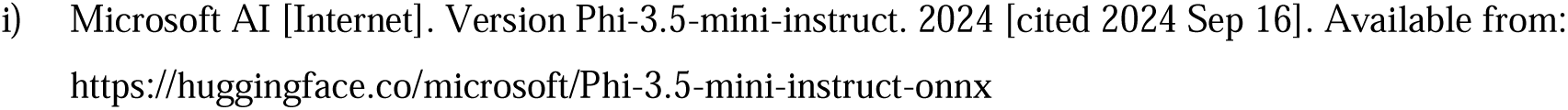
Characteristics of the large language models (LLMs) used in the study.

| Model | Version | Company | License | Study Tasks | Usage |
| --- | --- | --- | --- | --- | --- |
| GPT-4<sup>a</sup> | 128k turbo version<br>2024-04-09 | openAI | Commercial | Generate codes | API calls |
| GPT-3.5<sup>b</sup> | 16k turbo version<br>0125 | openAI | Commercial | Generate codes | API calls |
| Llama-3.1<sup>c</sup> | 70B-instruct | Meta | Open access | Generate codes,<br>Evaluate LLMs | Local GPU cluster |
| Llama-3.1<sup>d</sup> | 8B-instruct | Meta | Open access | Generate codes | Local GPU cluster |
| Qwen-2<sup>e</sup> | 72B-instruct | Alibaba Cloud | Open access | Generate codes,<br>Evaluate LLMs | Local GPU cluster |
| Qwen-2<sup>f</sup> | 7B-instruct | Alibaba Cloud | Open access | Generate codes | Local GPU cluster |
| Gemma-2<sup>g</sup> | 27B-it | Google | Open access | Generate codes | Local GPU cluster |
| Gemma-2<sup>h</sup> | 9B-it | Google | Open access | Generate codes | Local GPU cluster |
| Phi-3.5<sup>i</sup> | mini-instruct | Microsoft | Open access | Generate codes | Local GPU cluster |

- a) ChatGPT [Internet]. Version 4. 2024 [cited 2024 Sep 16]. Available from: https://www.openai.com/chatgpt - b) OpenAI. ChatGPT-3.5 Turbo [Internet]. Version 3.5. 2024 [cited 2024 Sep 16]. Available from: https://openai.com/index/gpt-3-5-turbo-fine-tuning-and-api-updates/ - c) Meta AI [Internet]. Version Llama 3.1-70B. 2024 [cited 2024 Sep 16]. Available from: https://www.llama.com/ - d) Meta AI [Internet]. Version Llama 3.1-8B. 2024 [cited 2024 Sep 16]. Available from: https://www.llama.com/ - e) Alibaba AI [Internet]. Version Qwen-2-72B-Instruct. 2024 [cited 2024 Sep 16]. Available from: https://www.alibabacloud.com/en/solutions/generative-ai/qwen?_p_lc=1 - f) Alibaba AI [Internet]. Version Qwen-2-7B-Instruct. 2024 [cited 2024 Sep 16]. Available from: https://www.alibabacloud.com/en/solutions/generative-ai/qwen?_p_lc=1 - g) Google AI [Internet]. Version Gemma-2-27b-it. 2024 [cited 2024 Sep 16]. Available from: https://ai.google.dev/gemma - h) Google AI [Internet]. Version Gemma-2-9b-it. 2024 [cited 2024 Sep 16]. Available from: https://ai.google.dev/
- i) Microsoft AI [Internet]. Version Phi-3.5-mini-instruct. 2024 [cited 2024 Sep 16]. Available from: https://huggingface.co/microsoft/Phi-3.5-mini-instruct-onnx

### Retrieval Augmented Generation Setup

To create code description embeddings, the GIST-large-embedding-v0 model was utilized, as it has been identified as effective for clinical tasks in a previous study. ^11^ The model embedded all ICD descriptions assigned across MSHS EDs through 2023. Using the Facebook AI Similarity Search (FAISS) library ^12^, descriptions were indexed for rapid retrieval. This enabled a search for the top 10 similar entries (via cosine similarity of code descriptions). Retrieved entries included the similar code description, its associated code, and the assignment frequency during 2021-2023.

### Statical Analysis

We used descriptive statistics to summarize patient demographics and clinical variables. We initially assessed the rate of JSON failures for the first prompt per model. Following this, we evaluated the capability of each model to generate valid codes from a zero-shot prompt by comparing the produced codes with 2023 ICD-10-CM published codes. Furthermore, the production of “exact match” codes—defined as complete congruence between the provider-assigned code and the LLM-generated code—was assessed using both zero-shot and RAG-enhanced prompts for each model.

The comparison between LLM-generated and provider-assigned codes was quantified through head-to-head percentages of reviewers’ assigned accuracy and specificity “wins” to either LLM or provider codes. Both two LLMs (Llama-3.1, Qwen-2) and four physicians (two ED attendings and two ED residents) served as judges. We performed a qualitative analysis of GPT-4 generated codes using Llama-3.1’s judgment.

We analyzed Inter-rater reliability of the reviewers with Fleiss’ kappa. Chi-square tests were used to statistically compare the performance of LLMs against the provider’s gold standard, with a p-value of less than 0.05 indicating statistical significance. All analyses were conducted using Python, version 3.9.17.

## Results

### Dataset Characteristics

Between January 2024 and April 2024, there were 81,723 ED visits which resulted in a home discharge. From this group, we randomly selected 500 patient visits for our study sample. The 500-patient sample had 316 unique ICD-10-CM codes (Supp. Figure S1). Table 2 presents the characteristics of the sample cohort. The database used for RAG, covering the years 2021 to 2023, contained 1,038,066 ED visits. This database featured 10,876 unique ICD-10-CM codes (Supp. Figure S2).

**Table 2.** Characteristics of the sample cohort.

| Variable | Details |
| --- | --- |
| Sex | Female: 271 (54.2%), Male: 229 (45.8%) |
| Race | Other: 247 (49.4%), Black: 147 (29.4%), White: 106 (21.2%) |
| Age | Median: 43.73 years (IQR*: 31.15 - 62.46) |
| Top five originally assigned ICD-10-CM Descriptions | Chest pain, unspecified (20); Nausea with vomiting, unspecified (12); Unspecified abdominal pain (9); Unspecified fall, initial encounter (9); Viral infection, unspecified (9) |

\* IQR- Interquartile range

### Prompt Response Failures

Our analysis of JSON failures during the initial prompt for assigning ICD codes and descriptions revealed significant variations among models. Figure 2 illustrates the count of failed attempts for each model to generate the required JSON format with "code" and "description" fields. GPT-4, Qwen-2-7B, llama-3.1-70B, and gemma-2-9b showed no failures, achieving a perfect success rate. In contrast, Phi-3.5-mini exhibited the highest number of failures at 85.

**Figure 2.**
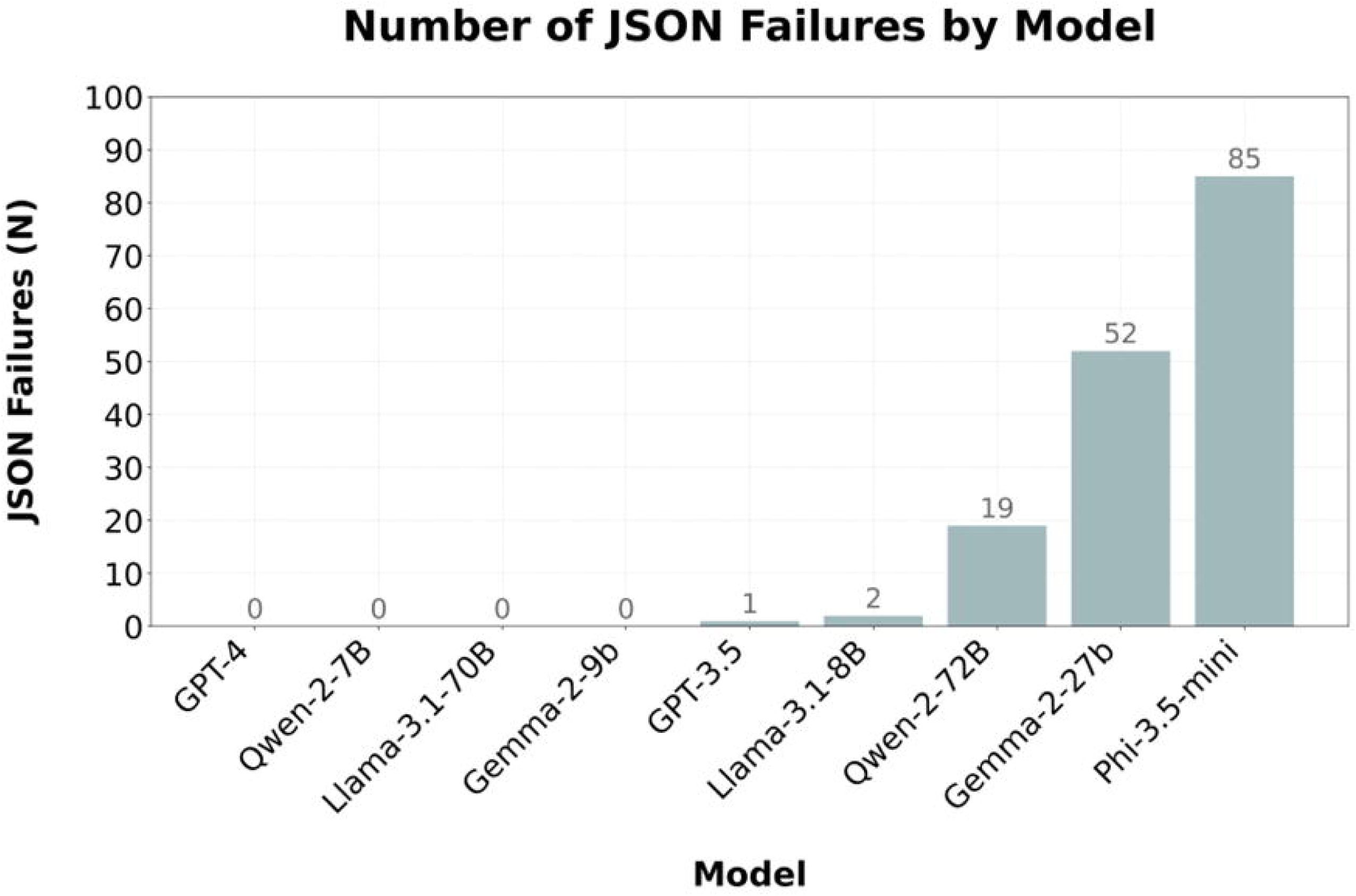
Number of JSON failures by model for the initial ICD code and description assignment prompt. Comparison number (N) of JSON format failures across different language models during the initial coding attempt.

### Valid ICD Code Generation

We evaluated the capability of the various LLMs to generate valid ICD codes from the ED notes (Figure 3a). The initial “zero-shot” prompt demonstrated varied performance across models, with valid codes ranging from a low of 42.4% (Qwen-2-7B) to a high of 95% (GPT-4). When the models were aided by the RAG-enhanced prompt, which constrained them to select from ten RAG-retrieved codes, the rate of valid codes improved dramatically, approaching near-perfect results. Specifically, the least successful model in the zero-shot scenario, Qwen-2-7B, surged to a 99.8% success rate with the RAG prompt, while GPT-4 maintained a perfect 100% valid code generation. The small number of non-valid codes in the RAG system represents the few cases of reverting back to zero-shot codes when the RAG prompt JSON call failed three times.

**Figure 3.**
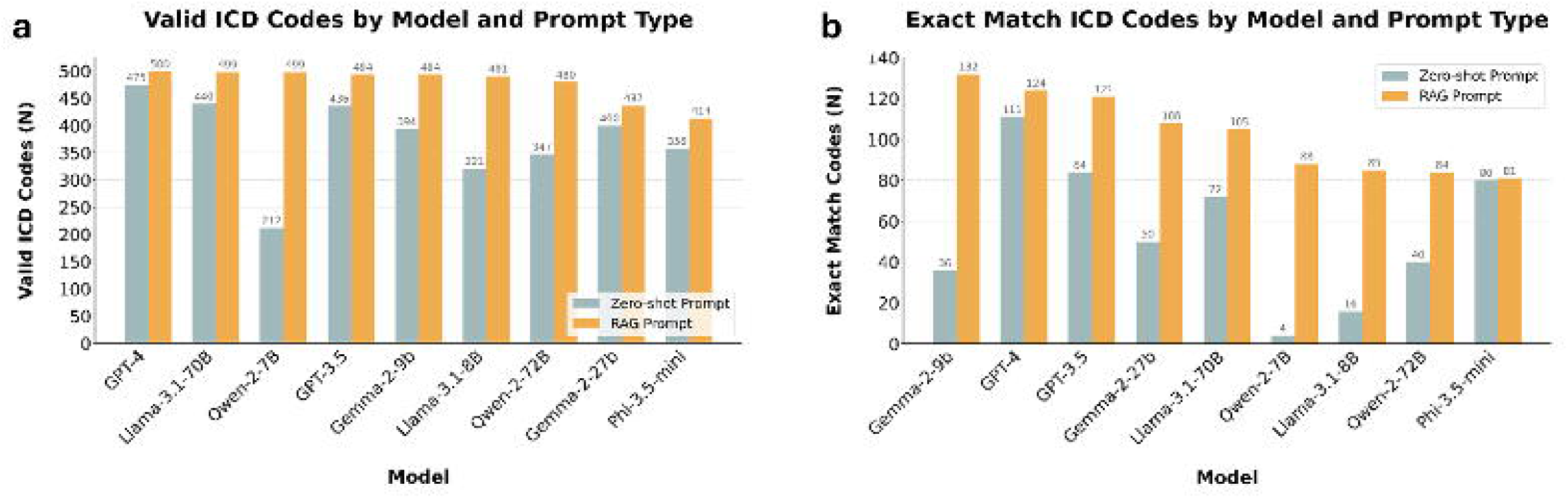
LLM Performance in ICD Code Generation from Clinical Notes. Percentage of valid ICD codes generated by each model. (B) Number of exact matches between LLM-generated and human-assigned ICD codes. Gray bars represent results from "zero-shot" prompting, while orange bars show results from RAG-enhanced prompting.

### Exact Match ICD Code Generation

Next, we analyzed the number of “exact matches” between human-assigned and LLM-assigned codes (Figure 3b). Across all models, the introduction of the RAG data led to an increase in the number of exact matches, though the extent of improvement varied markedly between models. For instance, the Qwen-2-7B model displayed a dramatic improvement, with exact matches increasing from a mere 4 (0.8%) in the zero-shot scenario to 88 (17.6%) with the RAG prompt. While the gemma-2-9b-it model jumped from 36 (7.2%) to 132 (26.4%), reflecting more than a threefold increase. In contrast, other models like GPT-4 model saw its exact matches rise from 111 (22.2%) to 124 (24.8%), and Phi-3.5 showed even more modest improvements, moving from 80 (16%) to 81 (16.2%).

### LLMs Evaluation of LLMs vs. Human-Assigned ICD Codes

In the study, two LLMs, Llama-3.1-70B, and Qwen-2-72B, served as judges to determine which coding— original human coding or LLMs assigned coding—was more accurate and specific. We excluded from the analysis cases in which the LLM and providers had the exact same codes.

### LLM Reviewers Inter-Rater Reliability

The inter-rater reliability between Llama and Qwen in their evaluations revealed a Fleiss’ Kappa value of 0.595 for accuracy and 0.688 for specificity.

### Accuracy

The assessment of LLMs for coding accuracy is presented in Figures 4a and 4b. Across the board, LLMs were consistently preferred over human-assigned codes, emphasizing their enhanced accuracy in medical coding tasks (p<0.001 for all models).

**Figure 4.**
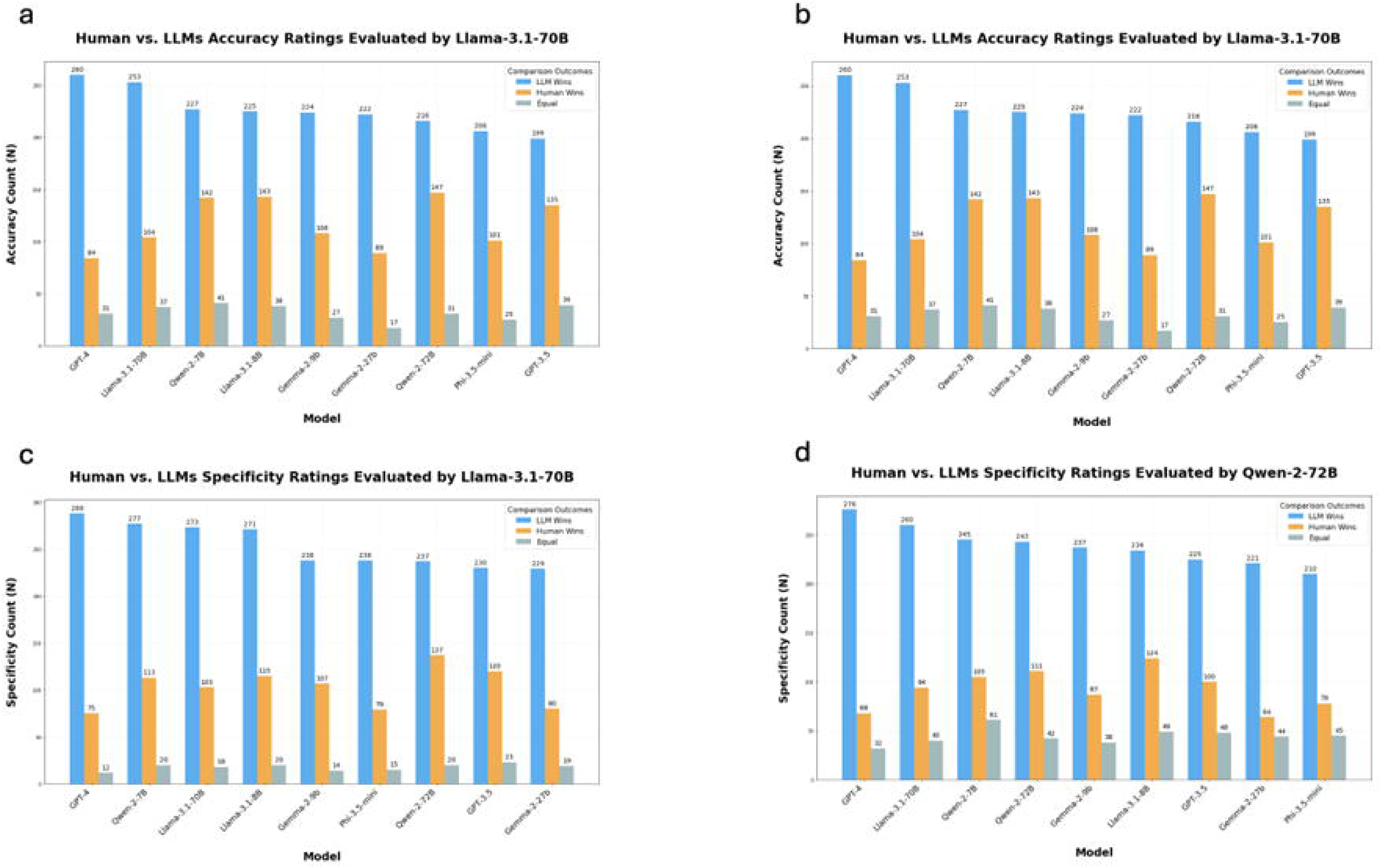
Comparative evaluations of ICD-10-CM code assignments: LLMs vs. Human coders. (a) This figure illustrates the evaluations made by Llama-3.1-70B comparing the accuracy of code assignments between all evaluated models and original human coders; (b) This figure illustrates the evaluations made by Qwen-2-72B comparing the accuracy of code assignments between all evaluated models and original human coders; (c) This figure illustrates the evaluations made by Llama-3.1-70B comparing the specificity of code assignments between all evaluated models and original human coders; (d) This figure illustrates the evaluations made by Qwen-2-72B comparing the specificity of code assignments between all evaluated models and original human coders.

The analysis revealed a notable performance by GPT-4, which stood out when evaluated by Llama-3.1-70B, achieving a higher preference with 260 instances where its coding was favored against 84 for human coding. Similarly, Llama-3.1-70B demonstrated superior coding skills when evaluated by Qwen-2-72B, with 218 instances of preference compared to 90 instances favoring human coders.

### Specificity

In the results of the specificity assessments conducted for ICD-10-CM coding, evaluations performed by Llama-3.1-70B and Qwen-2-72B reveal a significant preference for codes generated by LLMs over those assigned by humans (Figure 4c, 4d). These findings were consistent across several models, demonstrating the superior performance of LLMs in terms of specificity (p<0.001 for all models).

When evaluated by Llama-3.1-70B, GPT-4 was notably recognized for its precision, receiving the highest preference with 288 instances where its specificity was favored, compared to 75 where human coders were preferred. Both Qwen-2-7B and Llama-3.1-70B itself also showed strong results, with Llama-3.1 being preferred in 273 cases against 103 for human coders. This pattern underscores the robustness of LLMs in ensuring detailed and precise coding.

Similarly, under the evaluation of Qwen-2-72B, GPT-4 maintained its high performance, with 276 cases favoring its specificity compared to 68 for humans. Other models like Gemma-2-9b, although performing well, did not reach the high preference levels observed for GPT-4 or Llama-3.1-70B.

### Human Evaluation of GPT-4 vs. Human-Assigned ICD Codes

#### Human Reviewers Inter-Rater Reliability

In the evaluation of human reviewers’ inter-rater reliability for the criteria of accuracy and specificity, Fleiss’ Kappa was utilized. The calculated Fleiss’ Kappa value for accuracy was 0.307. For specificity, the Fleiss’ Kappa value was slightly higher at 0.346.

#### Accuracy

In a focused evaluation involving 200 random cases where GPT-4 and provider-assigned ICD codes did not exactly match, the human reviewers (two attendings and two residents) were blindly tasked with rating the accuracy of each assignment (Figure 5a). GPT-4 consistently demonstrated superior accuracy. GPT-4’s performance ranged from 1.23 to 2.69 times better than providers, with the largest difference observed with Resident 1 (129 for GPT-4 vs. 48 for human). Overall, reviewers preferred GPT-4 for accuracy in 447 cases, compared to 277 cases where human coders were preferred (p<0.001).

**Figure 5.**
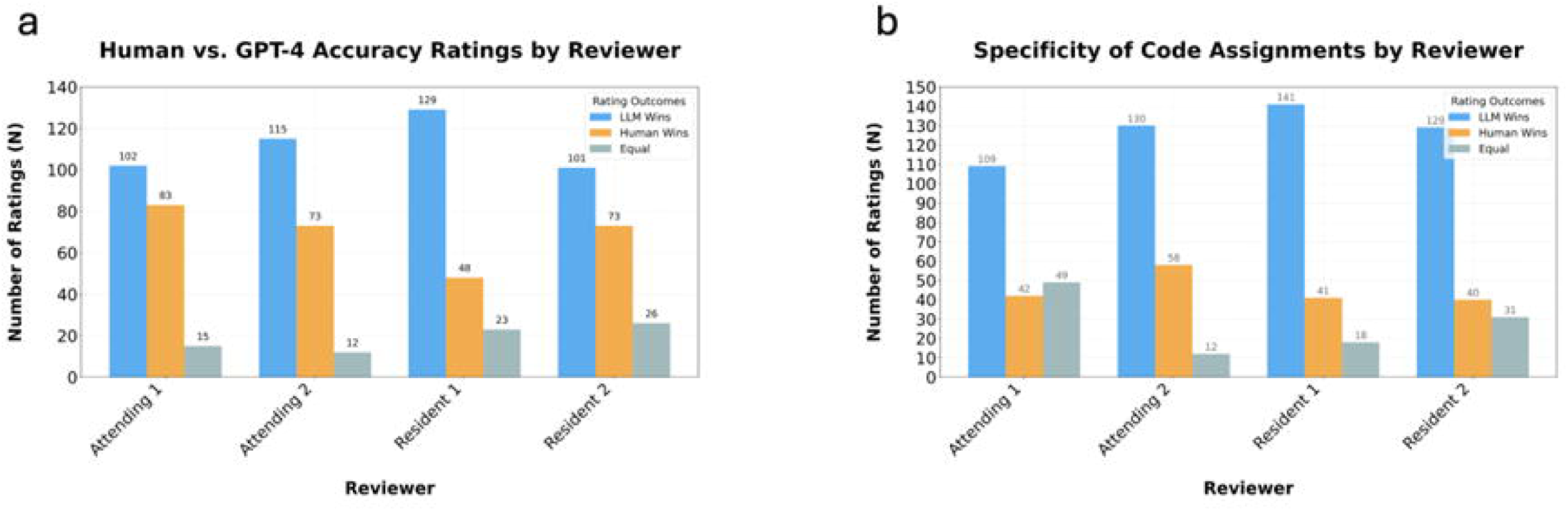
Human reviewers’ evaluation of GPT-4 vs. Provider-assigned ICD-10-CM codes. (a) This figure illustrates the evaluations made by two attending physicians and two residents comparing the accuracy of code assignments between GPT-4 and original human coders; (b) This figure displays the assessments conducted by two attending physicians and two residents evaluating the specificity of code assignments between GPT-4 and original human coders.

#### Specificity

The results of the physicians’ evaluation of code assignment specificity of GPT-4 vs. provider coders are presented in Figure 5b. The AI model, GPT-4, was notably favored, with GPT-4 codes being chosen 2.24 to 3.44 times more often than human codes. Overall, the reviewers preferred GPT-4 for specificity 509 times, compared to preferring human coders only 181 times (p<0.001).

#### Qualitative Analysis

Table 3 provides a comparison of the first 20 ICD-10-CM codes generated by GPT-4 against provider-assigned codes, evaluated by Llama-3.1 for both accuracy and specificity. Each entry includes cases where Llama-3.1 provided specific reasoning for its evaluations. In the comparative analysis of ICD-10-CM coding, GPT-4 consistently outperformed human-assigned codes. The model connected specific symptoms to precise diagnoses, such as associating epigastric pain with alcoholic gastritis.

**Table 3.**
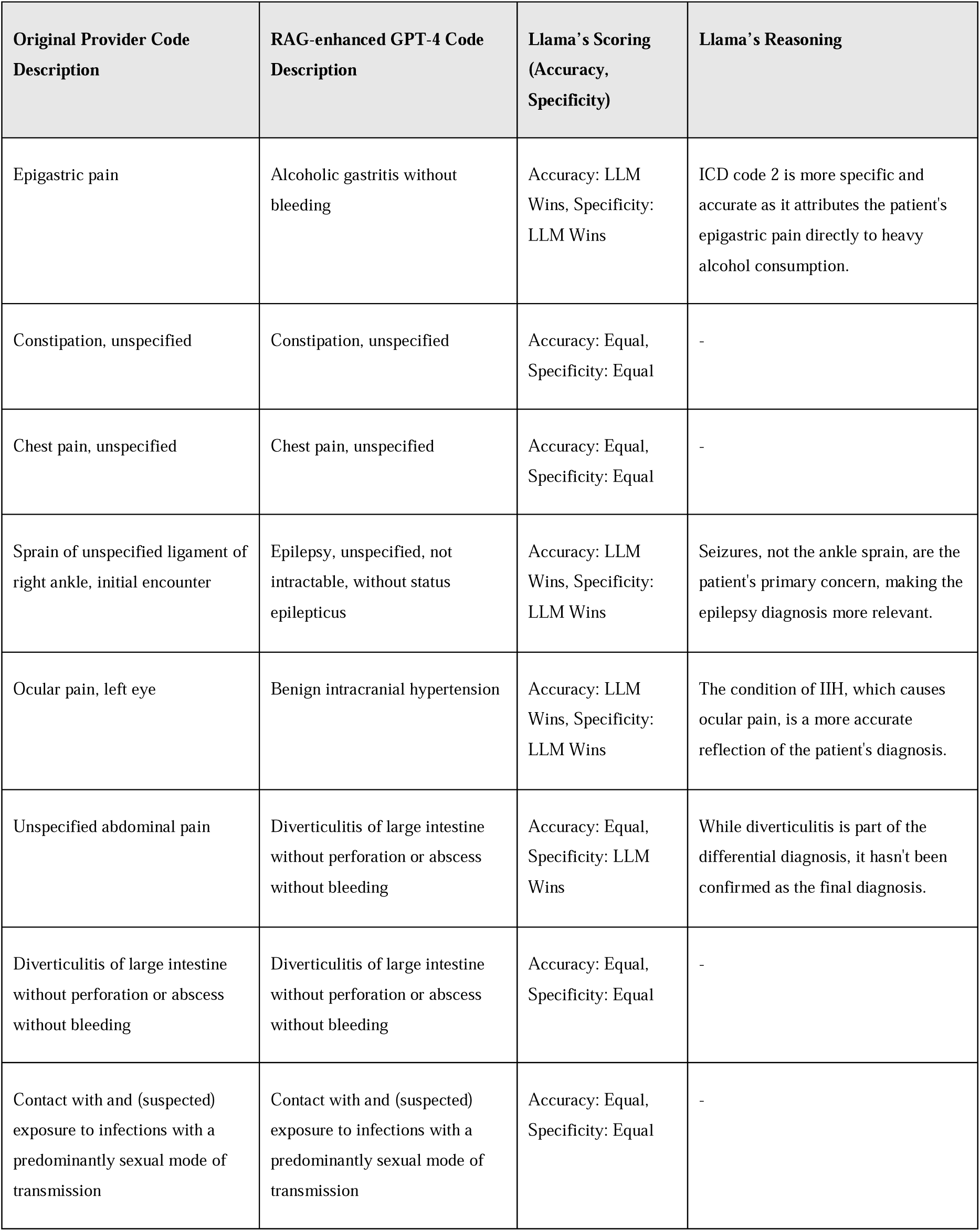
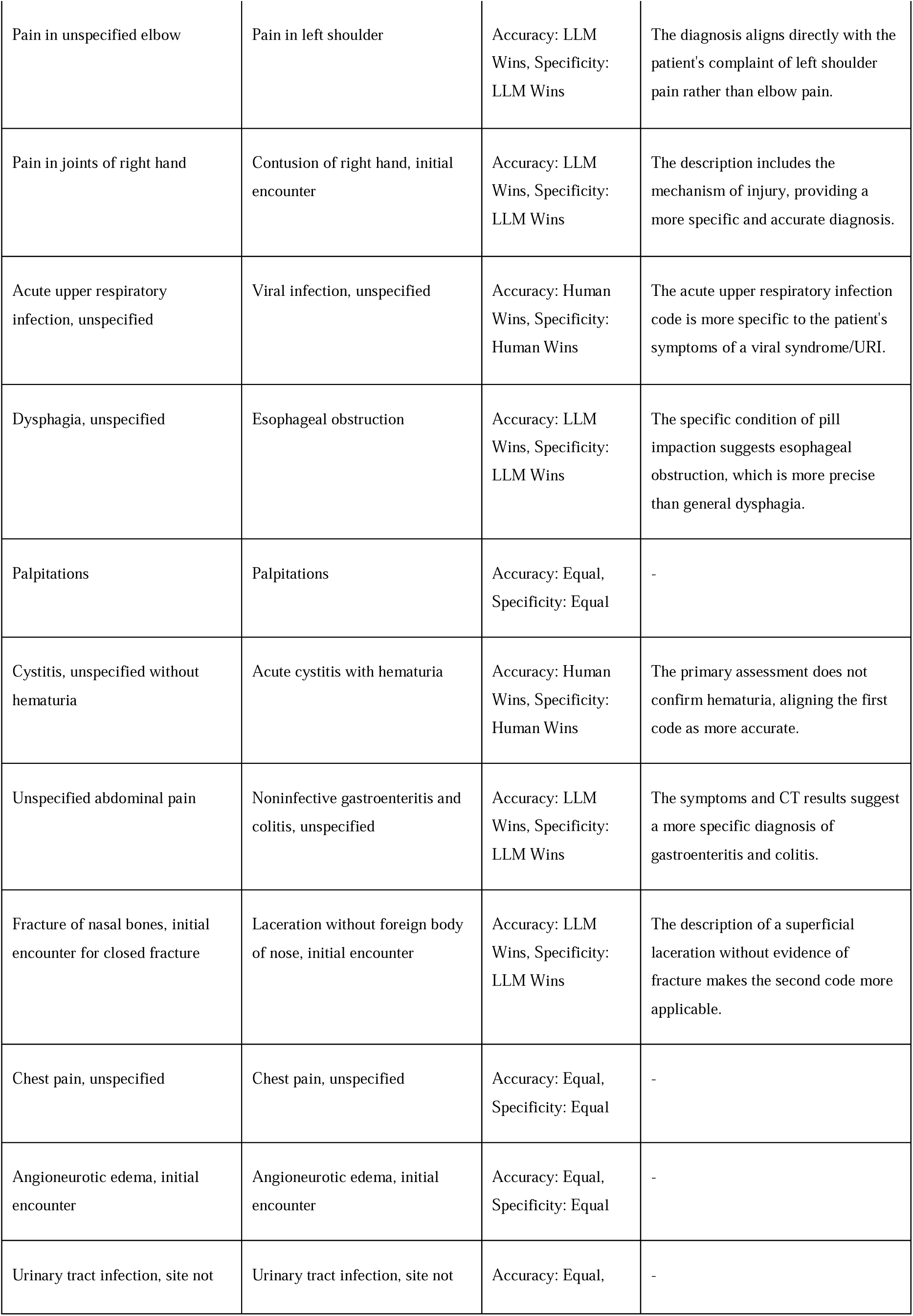
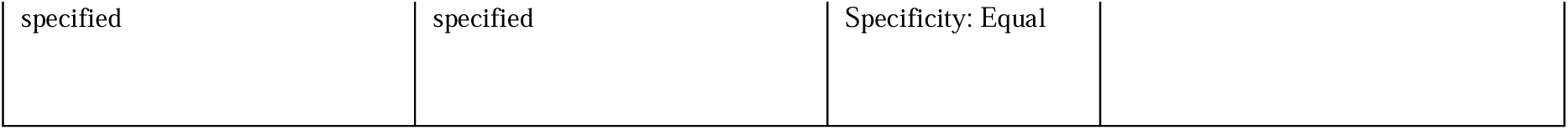
Comparison of the first 20 ICD-10-CM codes generated by GPT-4 against provider-assigned codes.

## Discussion

Medical coding is essential in clinical documentation, billing, reimbursement, and research, but manual processes are prone to errors and inconsistencies, particularly in fast-paced environments where documentation can be complex and time-sensitive. ^13^ This study investigates the application of the leading large language models (LLMs) enhanced with Retrieval-Augmented Generation (RAG) derived from real-world data for medical coding in ED settings. The study was conducted in a large, diverse, multi-site health system. The findings show that RAG-enhanced models significantly improve the specificity and accuracy of medical coding compared to traditional methods of provider documentation. Our results demonstrate that integrating RAG in real-time improves coding accuracy for LLMs, superior to manual human providers.

We show the importance of a RAG-enhanced LLM model in cases that involve complex clinical language and ambiguous documentation, which is typical in ED settings. Prior research has shown that manual coding is especially prone to error in situations where clinical complexity is high. ^14,15^ By retrieving relevant contextual information, the model enhances accuracy, even in cases where manual coders may struggle to identify the appropriate codes.

By utilizing real-time retrieval, such a model can perform better than existing provider-dependent systems at identifying accurate codes for conditions that evolve during patient care. ^16^ This aligns with existing literature that shows AI’s potential to transform clinical workflows by reducing administrative burdens while maintaining high accuracy. ^17^ As AI continues to show potential in becoming a part of clinical documentation workflows ^18^, this study demonstrates that, as of now, LLMs with RAGs are necessary for coding accuracy and efficiency, especially in high-stakes environments.

In our study, the performance increase of smaller open-access language models (7-8B parameters) when enhanced with RAG is particularly noteworthy. These models, despite their smaller size, achieve high levels of accuracy and specificity, broadly comparable to their larger counterparts. This finding underscores the efficiency of RAG in bolstering model performance without the need for extensive computational resources. ^19,20^ The scalability of smaller models implies broader applicability across diverse healthcare settings, even those with limited IT infrastructure. The use of RAG highlights another pathway where LLM can mitigate disparity among low resource reigns. Consequently, future research might benefit from focusing on optimizing retrieval mechanisms and ensuring high-quality data inputs to maximize the efficacy of smaller models in clinical applications.

Beyond its contribution to coding accuracy, the described framework also has implications for improving hospital operations. Accurate ICD coding is critical for billing processes, insurance claims, and overall healthcare system efficiency. ^21^ The RAG-LLM model’s ability to increase the number of correct and specific codes can lead to more timely reimbursements and reduce the financial burden on healthcare facilities. Moreover, by automating routine coding tasks, the model frees up healthcare professionals to focus on more critical decision-making tasks, thereby potentially improving patient outcomes.^22^ Not having to put in any diagnosis codes would reduce the number of physician “clicks” and the cognitive burden of having to think about the best single code to describe the clinical data.

However, the study also highlights some limitations that must be addressed for broader implementation. First, the model’s performance could degrade over time if not continuously updated with new human-generated coding data. As medical practices evolve and new diseases or treatments emerge, models that are not retrained may produce inaccurate results, a problem seen in other AI applications in healthcare. ^23^ Another limitation concerns the ethical considerations of using AI in clinical settings. ^24^ As the model automates decision-making, it introduces a risk of “black box” decision-making ^25^, where the reasoning behind coding choices is not transparent to clinicians and payers. The most problematic here would be systematic errors that can’t be identified over coding. This is much harder to trace when it’s a “black box” model. However, as we have demonstrated, the models can explain the rationale behind their code assignment. It is essential to build trust between humans and AI systems, ensuring transparency in how codes are generated and allowing for human oversight in critical cases. ^26^ Another challenge is the generalizability of AI models across different healthcare settings.^27,28^ However, our proposed system seems generalizable, as other systems can build their RAG databases using their own prior human-assigned codes in a similar fashion.

Further research is required to understand how well the model performs in other healthcare settings and for other medical specialties, which may have more complex clinical and coding scenarios. However, in more complex and specialized disciplines, automatic coding may turn out to be simpler because the model is able to narrow quickly compared to the very broad emergency medicine differential. Additionally, integrating AI into existing hospital information systems requires significant investment in infrastructure and training for staff, which may present barriers to adoption in smaller healthcare facilities.^29^

In conclusion, this study provides compelling evidence of the efficacy of RAG-enhanced LLMs in improving ICD-10-CM coding within ED settings. This model not only enhances the specificity and accuracy of medical coding but also promises to reduce the administrative burden on healthcare providers, improve patient outcomes through better documentation of conditions, and enhance medical research by enabling more precise cohort selection. However, further studies are necessary to fully understand the model’s limitations, ensure its long-term adaptability, and explore its applicability across a broader range of clinical environments.

### Research in context panel

#### Evidence before this study

*W*e searched PubMed, Embase, and the Cochrane Library for publications published between the database’s foundation and April 2024 using the terms "artificial intelligence", "machine learning", "ICD-10-CM coding", and "emergency department". We discovered various papers investigating the application of artificial intelligence (AI) in medical coding, but none that specifically assessed the performance of Retrieval-Augmented Generation (RAG) improved large language models (LLMs) for ICD-10-CM coding in emergency departments. Previous research found that when supplied only with code descriptions, LLMs frequently make wrong mappings, resulting in incorrect or invented codes.

#### The added value of this study

This is the first study to assess the performance of RAG-enhanced LLMs in producing ICD-10-CM codes from emergency department clinical notes and comparing them to codes provided by physicians. We found that RAG-enhanced LLMs overcome human coders in terms of accuracy and specificity when assigning ICD-10-CM codes. In addition, smaller open-access models, when RAG-enhanced, exhibited comparable levels of accuracy and specificity to larger models.

#### Implications of all the available evidence

Our findings indicate that RAG-enhanced LLMs have the potential to improve ICD-10-CM coding accuracy in emergency departments, thereby lowering administrative load and increasing coding efficiency. The equivalent performance of smaller, RAG-enhanced models to bigger ones demonstrates the scalability and efficiency of this method. Future studies should focus on the practical use of these technologies, such as their integration into existing hospital systems and their influence on clinical workflows and patient outcomes.

## Data Availability

All data produced in the present study are available upon reasonable request to the authors

## Ethical Approval

The study utilized a random sample of patients from the Mount Sinai Health System (MSHS), spanning multiple hospital EDs. Institutional Review Board (IRB) approval was obtained, and informed consent was waived by the IRB committee.

## Authors and contributors

E.K, I.T, R.F, and G.N. conceptualized the study; E.K, I.T, D.A., E.A, M.A, A.M and A.S. contributed to methodology; E.K and I.T prepared original draft; A.S, A.C, D.R, J.M, N.G, and B.C contributed to study visualization; R.F and G.N. supervised the study, accessed and verified the underlying data reported in the manuscript; All authors had full access to all the data and they read and agreed to the published version of the manuscript.

## Data Sharing

The datasets used and/or analyzed during the current study are available from the corresponding author upon reasonable request.

## Funding

This work was supported in part through the computational and data resources and staff expertise provided by Scientific Computing and Data at the Icahn School of Medicine at Mount Sinai and supported by the Clinical and Translational Science Awards (CTSA) grant UL1TR004419 from the National Center for Advancing Translational Sciences. Research reported in this publication was also supported by the Office of Research Infrastructure of the National Institutes of Health under award number S10OD026880 and S10OD030463. The content is solely the responsibility of the authors and does not necessarily represent the official views of the National Institutes of Health. The funders played no role in study design, data collection, analysis and interpretation of data, or the writing of this manuscript.

Declaration of generative AI and AI-assisted technologies in the writing process

During the preparation of this work the author(s) used [NAME TOOL / SERVICE] in order to [REASON]. After using this tool/service, the author(s) reviewed and edited the content as needed and take(s) full responsibility for the content of the publication.

## Supplementary Materials

**Prompt 1.** Initial ICD-10-CM Code Prediction Prompt.

**Prompt 2.** ICD-10-CM Code Refinement Prompt

**Prompt 3.** LLM Evaluation Prompt for Code Assignment Comparison

**Figure S1.** Top 10 Diagnoses by Percentage – Sample CCS

**Figure S2.** Top 10 Diagnoses by Percentage – Cohort CCS

