## Supplementary material for "Assessing Retrieval-Augmented Large Language Model Performance in Emergency Department ICD-10-CM Coding Compared to Human Coders": supp data

**Supplementary Materials**

**Prompt 1.** Initial ICD-10-CM Code Prediction Prompt.

**Prompt 2.** ICD-10-CM Code Refinement Prompt

**Prompt 3.** LLM Evaluation Prompt for Code Assignment Comparison

**Prompt 1. Initial ICD-10-CM Code Prediction Prompt**

Below are a patient's ED visit notes.

Determine the most likely ICD-10-CM ED primary diagnosis that was assigned in this case.

The description must match official 2023 ICD-10-CM descriptions.

Return the diagnosis in JSON format:

{{"description": "YOUR DIAGNOSIS DESCRIPTION", "code": "ICD-10 CODE"}}

\#### Patient Notes: {text}

**Prompt 2. ICD-10-CM Code Refinement Prompt**

Below are ED visit notes and a list of ICD-10-CM codes and descriptions.

Also included are the frequencies of these diagnoses in our ED.

From this list, choose the ICD-10-CM primary diagnosis that was most likely assigned in this case.

Return the diagnosis in JSON format:

{{"description": "YOUR DIAGNOSIS DESCRIPTION", "code": "ICD-10-CM CODE"}}.

\#### Patient Notes: {text}

**Prompt 3. LLM Evaluation Prompt for Code Assignment Comparison**

You are given patient ED notes and two ICD code descriptions.

Your task is to evaluate which ICD description is (1,2,equal):

more accurate - i.e., presents the cases without mistakes

more specific - i.e. presents the case with more specific details

Please return your evaluation in the following JSON format:

{{"reason": "<a short explanation for your decision>", "accuracy": <"1" OR "2" OR "equal">, "specificity": <"1" OR "2" OR "equal">}}

\#### Patient Notes: {text}

\#### ICD Code Descriptions:

1: {ICD1}

2: {ICD2}

**Figure S1: Top 10 Diagnoses by Percentage – Sample CCS**

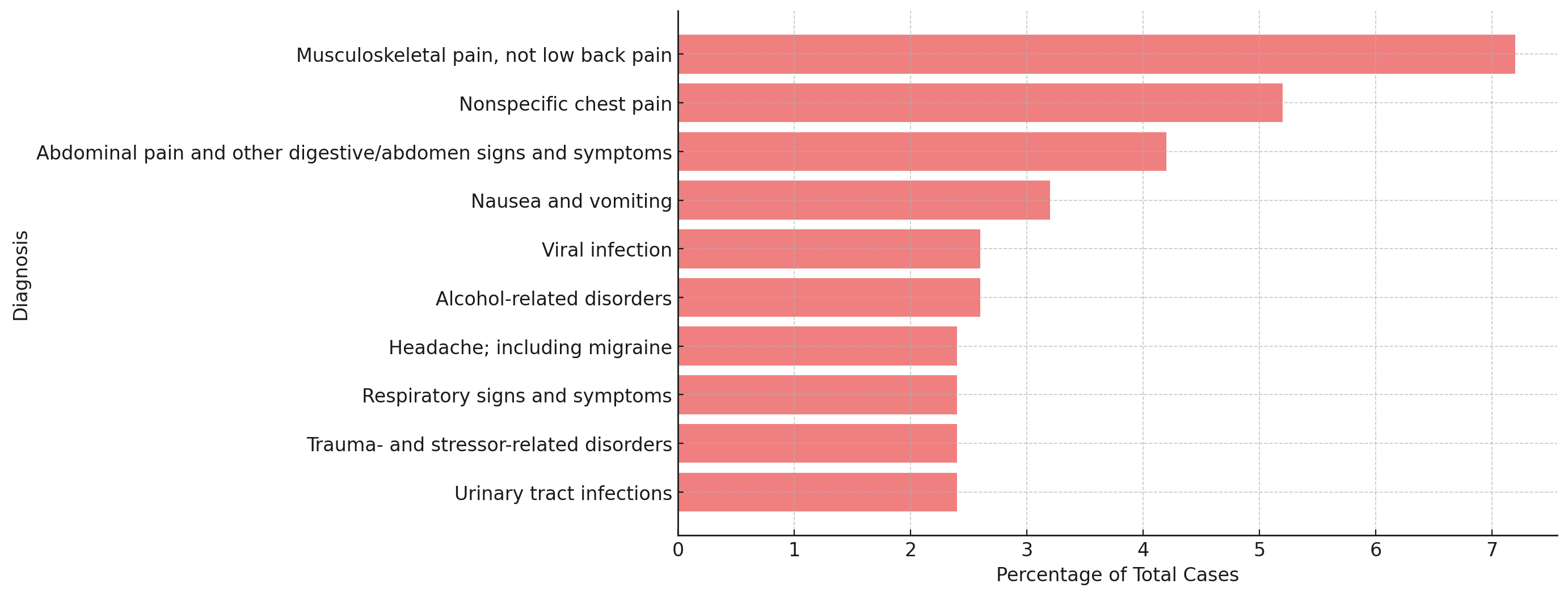

**Figure S2: Top 10 Diagnoses by Percentage – Cohort CCS**

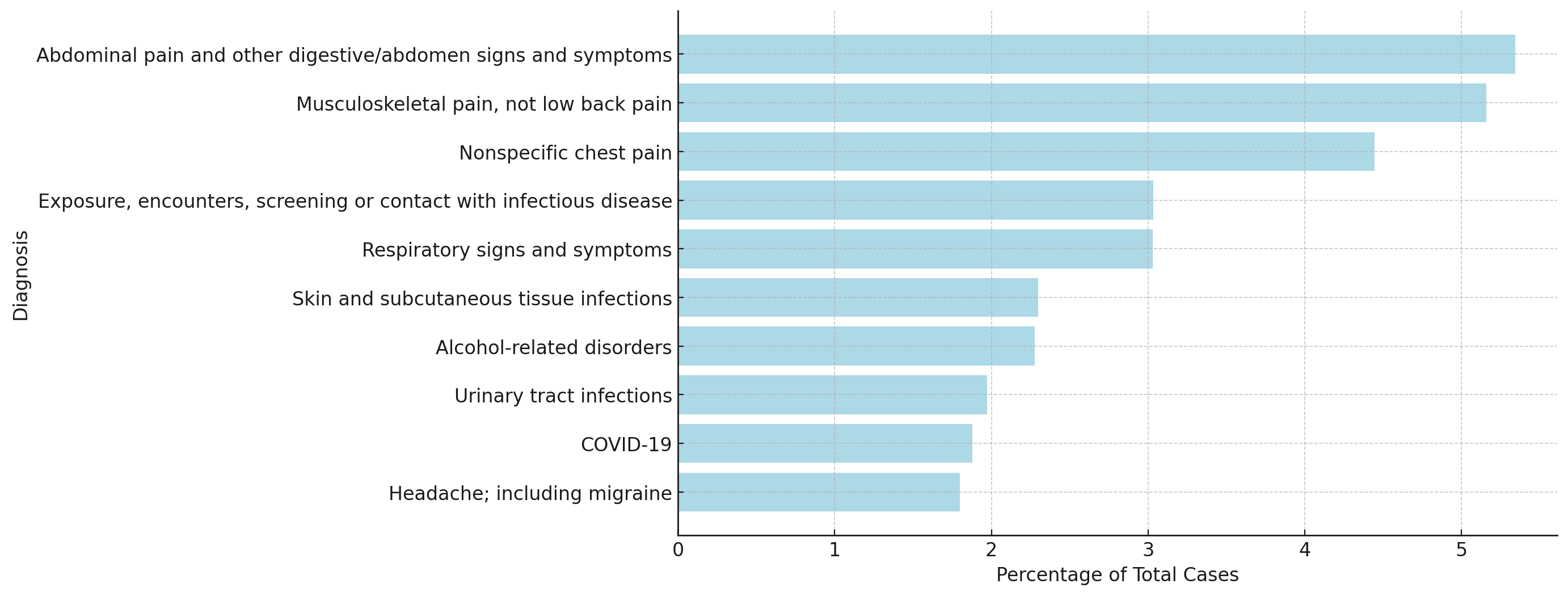
